# Direct Amplification-Free Nanopore Sequencing of *Leishmania* from Cutaneous Biopsies

**DOI:** 10.64898/2026.09.12.26362874

**Authors:** Faisal Alkhaldi, Abdullatif S. Al Rashed, Christian Frøkjær-Jensen

## Abstract

**Background:** PCR confirms cutaneous leishmaniasis but yields limited species data. Direct nanopore sequencing could offer broader genomic evidence without amplification, though the value of long-read sequencing depends on target recovery, accurate species assignment, and cost.

**Methods:** Sixteen biopsy-DNA extracts (15 PCR-positive, one negative) were sequenced without target amplification using 16-plex adaptive human depletion. Three extracts were additionally analyzed via standard sequencing and adaptive target enrichment. A positive call required at least three nuclear *Leishmania* reads spanning distinct 10-kb loci across at least two chromosomes. Species evidence was evaluated by competitive alignment, Kraken2, and markers.

**Results:** Fourteen PCR-positive biopsies met the detection rule (93.3% agreement) within 3.5 h of sequencing with the negative biopsy yielding no qualifying reads. Adaptive enrichment increased the target fraction 3.1–5.8-fold but recovered only 12–62% of the absolute target bases obtained by standard sequencing and did not shorten detection time. Unexpected *L. aethiopica* assignments motivated a held-out benchmark, revealing that classifiers systematically misassigned 97.1–98.3% of *L. tropica* fragments to *L. aethiopica* due to reference bias. Marker analysis provided a sparse but useful locus-specific check contrasting these assignments. Modeled sequencing costs (US$30–72/specimen) exceeded endpoint PCR (US$5).

**Conclusions:** PCR-free nanopore sequencing recovers conservative multi-locus *Leishmania* evidence directly from biopsies. Adaptive enrichment improved target fraction without improving absolute recovery, while incomplete reference representation limited species interpretation even when genome-wide classifiers agreed. The current approach is therefore better positioned as a genomic adjunct for surveillance settings than as a replacement for routine PCR.

## 1 Introduction

Leishmaniasis comprises a spectrum of vector-borne diseases caused by protozoan parasites belonging to the genus *Leishmania*. Cutaneous leishmaniasis is the most common clinical form and can produce persistent, disfiguring lesions, yet its clinical appearance frequently overlaps with other infectious and inflammatory conditions [1, 2]. Laboratory confirmation commonly combines microscopy, culture, histopathology, and nucleic acid amplification. PCR is generally more sensitive than microscopy or culture, but targeted assays report only the target region interval selected in advance and often lack robust species resolution [2, 3].

Species identification is critical for epidemiology, prognosis, and treatment selection, particularly where distinct *Leishmania* species co-circulate [2, 4]. Conventional typing methods target loci such as ITS1, ITS2, the miniexon, *HSP70*, *HSP20*, and cytochrome *b*; however, discriminatory power varies across species complexes and geographic populations [5–11]. Genome plasticity, population structure, hybridization, and unevenly sampled reference genomes further complicate the translation of sequence similarity into definitive species assignments [12–16].

Nanopore sequencing provides long reads and real-time data analysis during acquisition. Published *Leishmania* nanopore assays have largely sequenced LAMP, RPA, or PCR products, including *HSP70* amplicons [17–20]. Culture-independent parasite genomics from clinical specimens has been demonstrated using hybrid capture or selective whole-genome amplification [21–23], and untargeted short-read metagenomics has detected visceral leishmaniasis in clinical samples [24]. Direct, amplification-free long-read sequencing would preserve genome-wide context and eliminate primer bias, but the low fraction of parasite DNA in tissue extracts remains a major obstacle.

Nanopore adaptive sampling accepts or ejects individual molecules after real-time comparison of their initial sequence against a user-defined reference. The approach can enrich low-abundance organisms or deplete host DNA without additional wet-laboratory capture [25–27]. However, an increase in target *fraction* does not necessarily translate to more target bases or earlier detection because molecule length, pore occupancy, rejection overhead, and run-specific yield all contribute to absolute recovery [26, 28, 29].

Here, we evaluated PCR-free nanopore sequencing of biopsy-DNA extracts using a conservative, multi-locus nuclear-read rule for genus-level detection. We then addressed three linked questions: whether adaptive sampling improved target recovery and sequencing-only detection time; whether genome-wide classifiers could separate clinically relevant species when assemblies were withheld from their reference panels; and whether long reads spanning established markers supplied additional species evidence. Crucially, this work represents an analytical feasibility assessment rather than an estimate of clinical diagnostic accuracy.

## 2 Results

### 2.1 PCR-free sequencing enabled genus detection and species analysis

The dataset comprised 16 biopsy-DNA extracts: 15 positive and one negative by PCR targeting the ribosomal internal transcribed spacer (ITS) (Figure 1A). All 16 extracts were sequenced in a multiplexed adaptive human-depletion run. A subset of three extracts were additionally sequenced under standard and adaptive *Leishmania* target-enrichment conditions.

**Figure 1:**
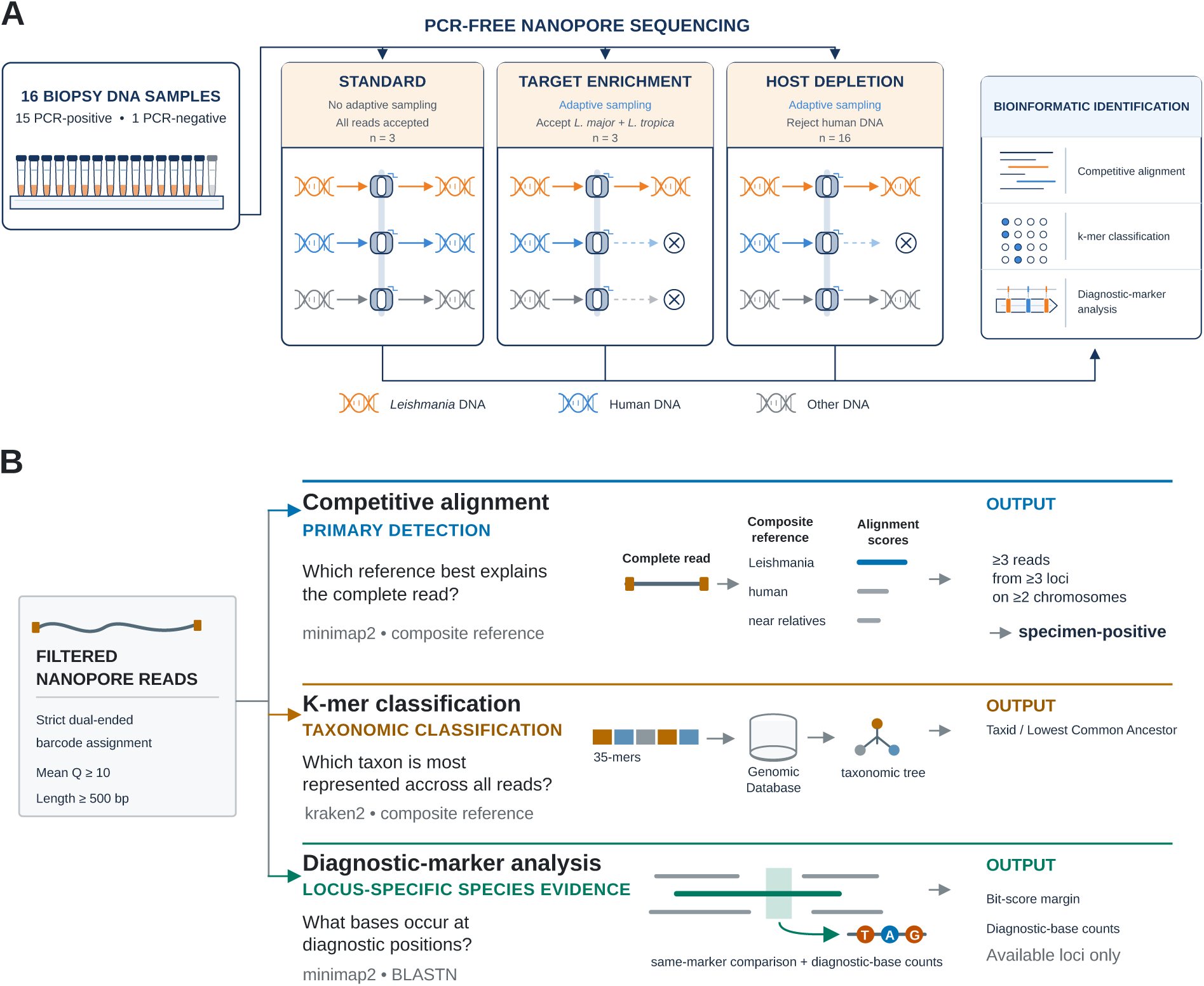
Experimental design and bioinformatic workflow for PCR-free nanopore detection of *Leishmania*. (A) Schematic illustrating the study design and three sequencing modes. Sixteen biopsy-DNA specimens (15 PCR-positive and one PCR-negative) were included in an adaptive host-depletion run, while three specimens were also sequenced using standard sequencing and adaptive target enrichment. Standard sequencing accepted all molecules, target enrichment retained *Leishmania*-matching molecules, and host depletion rejected human-matching molecules. Orange, blue, and gray represent *Leishmania*, human, and other DNA, respectively. (B) Overview of the three complementary strategies used to analyze demultiplexed, quality-filtered reads. Competitive alignment was used for conservative genus-level detection, Kraken2 for genome-wide taxonomic classification, and diagnostic-marker analysis for locus-specific species identification.

Reads first underwent stringent dual-ended demultiplexing, followed by filtering for a mean Phred score of 10 and a minimum length of 500 bp. The primary genus-detection analysis competitively aligned each complete read against *Leishmania*, human, near-relative, and contaminant references. A specimen met the sequencing-positive rule only if it contained at least three qualifying reads mapping to three distinct 10-kb loci distributed across at least two chromosomes. Genome-wide *k*-mer classification and marker-specific analysis were used to provide complementary taxonomic evidence (Figure 1B).

### 2.2 Multi-locus reads detected 14 of 15 PCR-positive biopsies

Fourteen of 15 PCR-positive biopsies met the multi-locus sequencing rule, corresponding to a positive percent agreement of 93.3% (exact 95% confidence interval, 68.1–99.8%; Figure 2A). The 14 detected specimens contained 12–2,470 qualifying reads. The remaining PCR-positive specimen contained only one qualifying read and therefore did not meet either the read-count or multi-locus components of the rule. The sole PCR-negative biopsy contained no qualifying reads. Because the comparator-negative group comprised only one specimen, this observation was reported descriptively and no specificity estimate was calculated.

**Figure 2:**
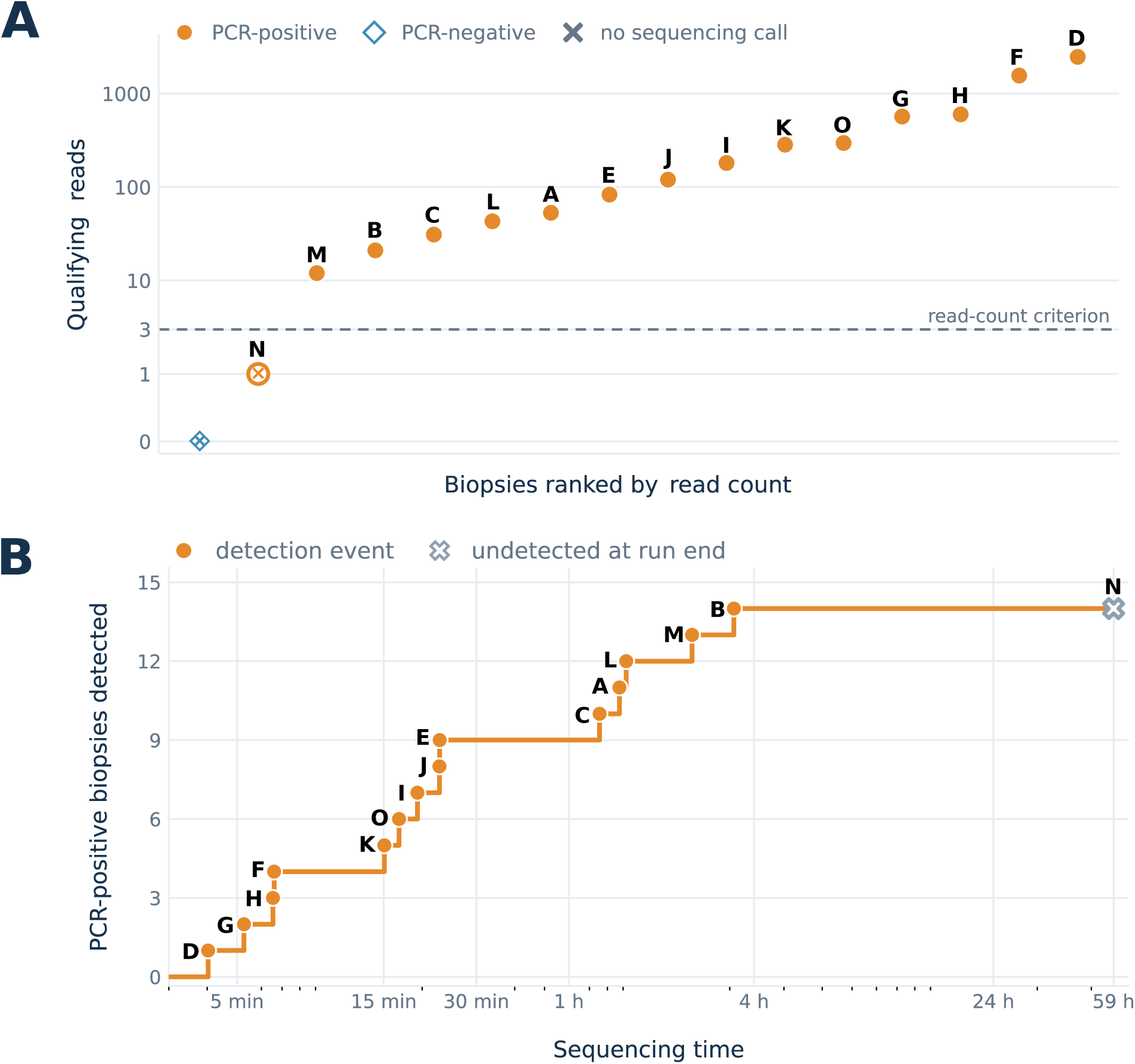
Direct detection of *Leishmania* in biopsy DNA and sequencing-only time to detection. (A) Qualifying nuclear *Leishmania* reads recovered from each biopsy, ordered by read count. Orange circles represent PCR-positive specimens, the blue diamond represents the PCR-negative specimen, and crosses mark specimens that did not meet the sequencing-positive rule. The dashed line indicates the three-read threshold; a positive call also required reads from three distinct 10-kb loci across at least two chromosomes. The y-axis uses a symmetric-log scale to display zero values. (B) Cumulative number of PCR-positive biopsies meeting the multi-locus detection rule as reads were replayed chronologically. Orange points indicate individual detection events, and the cross marks the specimen that remained undetected at the end of the run. Time is measured from the start of sequence acquisition and excludes sample preparation, basecalling, and computational analysis.

Chronological replay of read acquisition showed that threshold crossing occurred throughout the early portion of the run (Figure 2B). The first detected specimen met the rule after approximately 4 min of sequencing, and all 14 sequencing-positive specimens had crossed it by approximately 3.5 h. These intervals measure sequence acquisition only; they do not include specimen processing, library preparation, basecalling, analysis latency, interpretation, or reporting.

### 2.3 Adaptive sampling increased target fraction without increasing absolute recovery

Adaptive target enrichment increased the target proportion in the three-extract technical comparison (Figure 3A). After using one-end specimen assignment to avoid loss of the distal barcode from ejected reads, the fraction of primary-filter sequence represented by qualifying nuclear *Leishmania* bases was 3.1–5.8 times higher under enrichment than under standard sequencing. Absolute recovery showed the opposite pattern: enrichment yielded only 12.0–62.0% as many qualifying target bases as the corresponding standard-run sequencing (Figure 3B).

**Figure 3:**
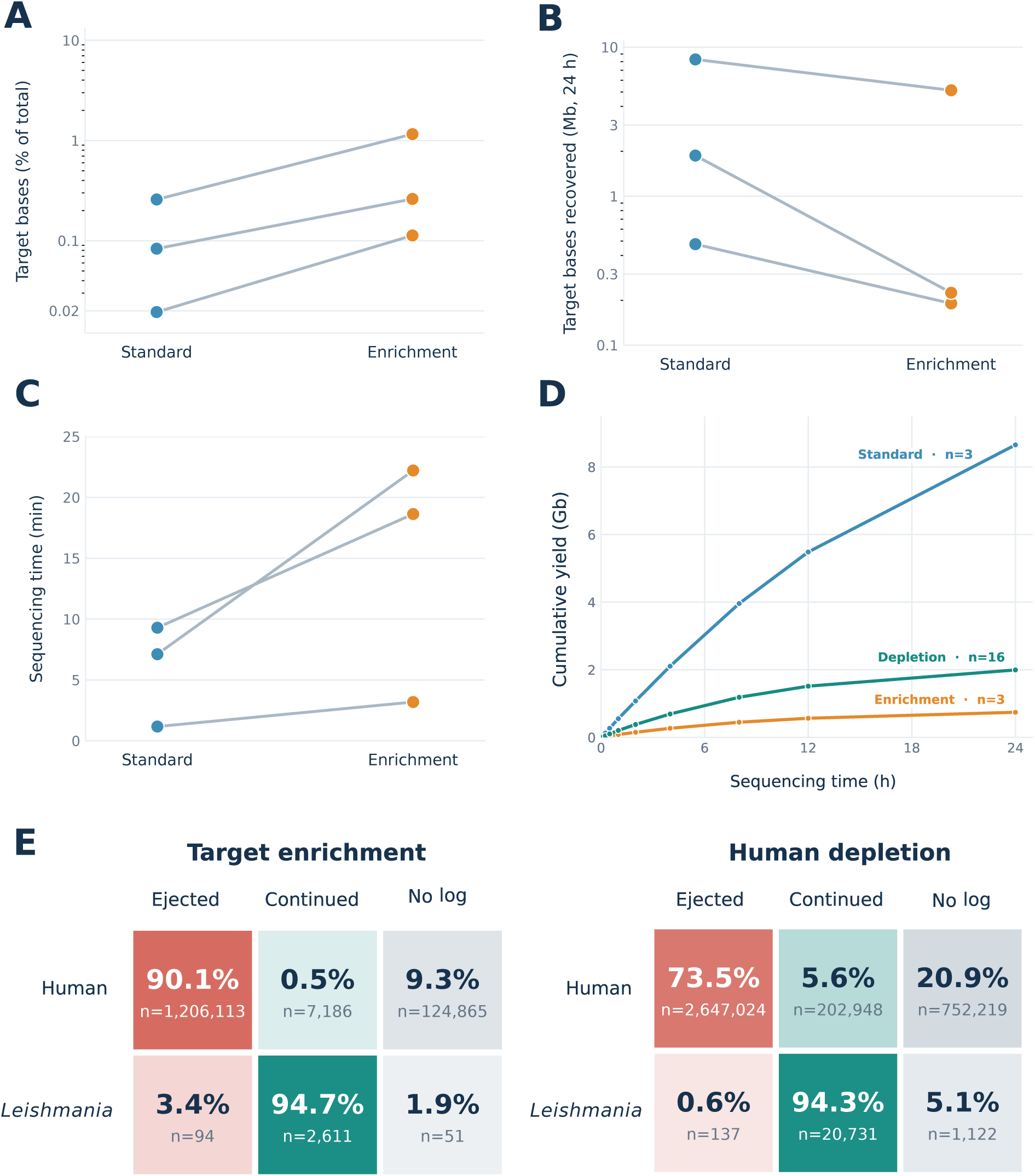
Target recovery and sequencing performance under adaptive sampling. (A) Proportion of one-end-assigned primary-filter bases assigned as qualifying nuclear *Leishmania* sequence during the first 24 h of standard and adaptive target-enrichment sequencing. (B) Absolute number of qualifying nuclear *Leishmania* bases recovered over the same period. Both y-axes are logarithmic. (C) Sequencing time required to meet the genus-level detection rule for the three paired extracts. (D) Cumulative barcode-independent primary-filter yield during the first 24 h of the standard three-sample, target-enrichment three-sample, and host-depletion 16-sample runs. (E) Terminal adaptive-sampling decisions for reads classified offline as human or qualifying nuclear *Leishmania* sequence in the target-enrichment run and a separate host-depletion run.

The enrichment run did not shorten the time to detection. Detection times increased from 1.2 to 3.2 min, 7.1 to 22.2 min, and 9.3 to 18.6 min across the three paired extracts (Figure 3C). After 24 h, primary-filter yield was 8.65 Gb in the standard three-sample run, 0.742 Gb in the target-enrichment three-sample run, and 1.99 Gb in the 16-sample human-depletion run (Figure 3D). Because each mode was represented by a single run with different sample counts, these trajectories cannot distinguish adaptive mode from library, flow-cell, run-day, or other run-specific effects.

Terminal decision logs nevertheless showed that adaptive decisions were strongly directional (Figure 3E). Barcode-independent linkage of terminal decisions in the target-enrichment run showed 1,206,113 of 1,213,299 human reads with matched logs were ejected (99.41%), while 2,611 of 2,705 qualifying nuclear target reads continued (96.52%). In the human-depletion run, 2,647,024 of 2,849,972 human reads were ejected (92.88%), and 20,731 of 20,868 qualifying nuclear target reads continued (99.34%).

### 2.4 Genome-wide classifiers could not reliably distinguish *L. tropica* from *L. aethiopica*

Genome-wide species assignments were examined for the 14 biopsies that met the genus-level detection rule (Figure 4A). In most specimens, both competitive alignment and Kraken2 overwhelmingly supported *L. major*. Biopsies K and L were exceptions. Both classifiers assigned most of the genome-wide evidence from Biopsy K to *L. aethiopica*, whereas the classifiers disagreed for Biopsy L: competitive alignment favored *L. major*, while Kraken2 favored *L. aethiopica*.

**Figure 4:**
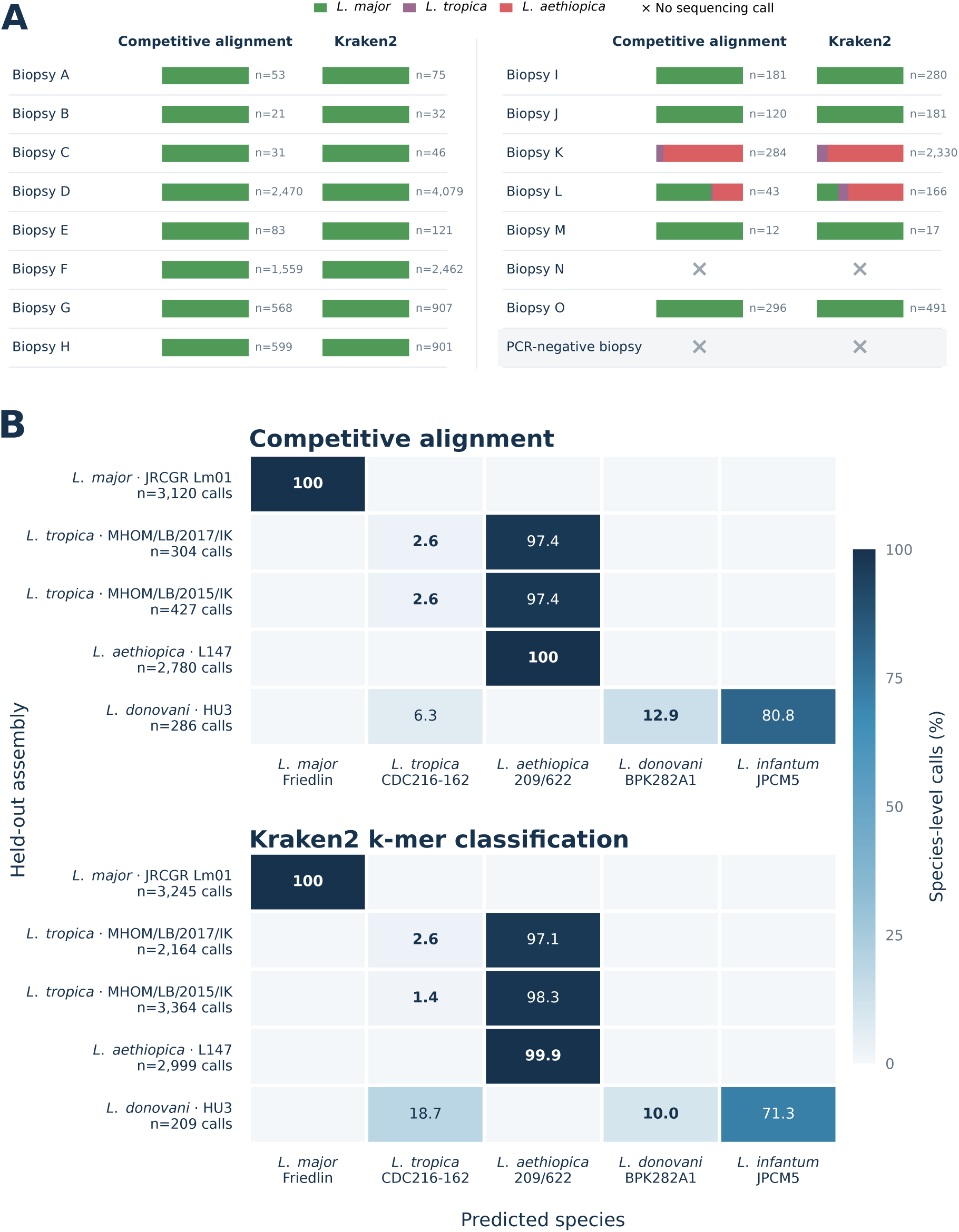
Genome-wide species assignment in clinical biopsies and held-out assemblies. (A) Relative support for *L. major*, *L. tropica*, and *L. aethiopica* from competitive alignment and Kraken2 classification in the biopsy cohort. Each bar is normalized across the displayed species-supporting reads, with *n* indicating the corresponding read count. Crosses mark specimens that did not meet the genus-level sequencing-positive rule and were therefore not assigned a species composition. (B) Classifier performance on deterministic 3-kb fragments sampled at 10-kb intervals from five nuclear assemblies excluded from the classifier references. Rows indicate the held-out species and strain, while columns indicate the assigned species and the reference strain representing it. Cell values show the percentage of species-level calls.

The prominent *L. aethiopica* signal was epidemiologically unexpected because the biopsies originated from the Eastern Province of Saudi Arabia, where molecular studies have identified *L. major* as the predominant cause of cutaneous leishmaniasis and have only recently documented *L. tropica*, with no established local transmission of *L. aethiopica* [30]. This raised an important analytical question: did the *L. aethiopica* assignments reflect genuine species-level evidence, or were the genome-wide classifiers unable to distinguish closely related species using the available reference assemblies?

To test classifier separability directly, we generated deterministic, error-free pseudoreads by sampling 3-kb fragments at 10-kb intervals from five nuclear genome assemblies excluded from the classifier reference panels (Figure 4B). Because these fragments preserved the original assembly sequences and contained no simulated nanopore errors, any misclassification would reflect limitations in reference representation or classifier separability rather than sequencing error.

Both methods correctly assigned all or nearly all calls from the held-out *L. major* and *L. aethiopica* assemblies to their archived species. In contrast, the two independent held-out *L. tropica* assemblies were systematically assigned to *L. aethiopica*: 97.4% of competitive-alignment calls and 97.1%–98.3% of Kraken2 calls favored *L. aethiopica*. This confusion was asymmetric, because the held-out *L. aethiopica* assembly was classified correctly. A similar limitation was observed within the *L. donovani* complex, where most fragments from the held-out *L. donovani* assembly were assigned to *L. infantum* by both alignment (80.8%) and Kraken2 (71.3%).

These results demonstrate that, under the reference-panel design, genome-wide agreement between the two classifiers did not guarantee a correct species label. In particular, the available references were non-identifying for the tested *L. tropica* genomes and produced a systematic bias toward *L. aethiopica*. This likely reflects both the close evolutionary relationship between these species and the limited representation of their population-level genomic diversity [13, 15, 31]. Consequently, the genome-wide *L. aethiopica* assignments in Biopsies K and L could not be taken at face value and motivated an additional analysis focused on established species-informative markers.

### 2.5 Marker reads provided complementary species evidence in ambiguous biopsies

Because the held-out benchmark showed that genome-wide methods could misclassify *L. tropica* as *L. aethiopica* even when provided with error-free sequences, we next asked whether reads spanning established diagnostic markers could provide more interpretable, locus-specific species evidence. This analysis was therefore undertaken specifically to interrogate the unexpected genome-wide assignments, rather than as an independent species-classification exercise.

We first evaluated seven established markers in six public whole-genome nanopore runs with known species labels (Figure 5A). Resolution varied substantially among loci and species groups. ITS2 and cytochrome *b* each produced a plurality matching the archived species in five of six runs, whereas the *HSP70* coding sequence was unresolved in four runs. Miniexon, *HSP20*, and the *HSP70* 3*^′^* UTR also produced discordant pluralities in some closely related species groups. These differences were consistent with variation in pairwise divergence among the available marker references (Supplementary Figure S1). Notably, the *L. tropica* and *L. aethiopica HSP70*-coding references differed by a median of only two aligned bases, making this locus poorly suited to discriminating the two species.

**Figure 5:**
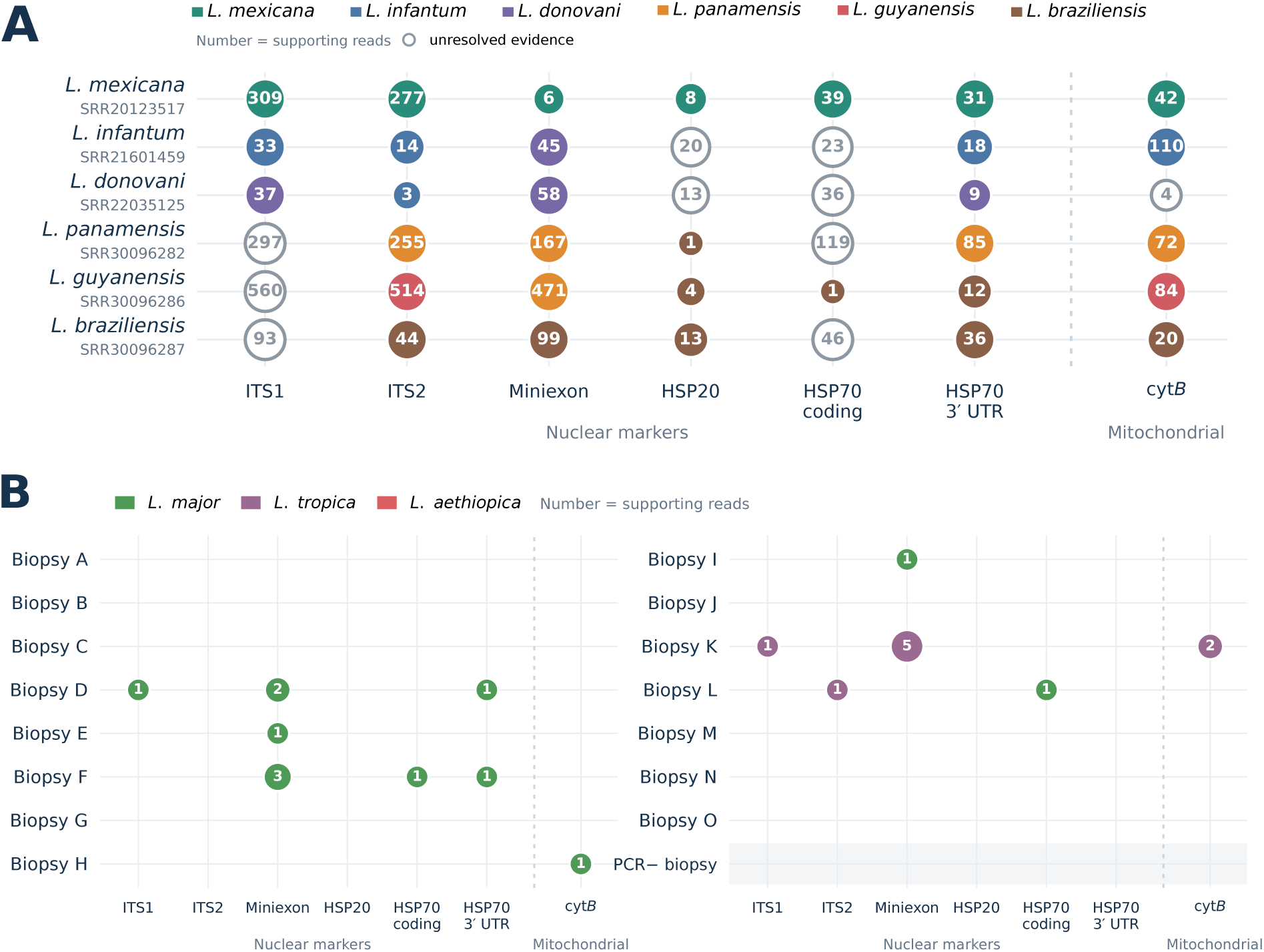
Marker-based species evidence in public nanopore datasets and clinical biopsies. (A) Species assignments across seven diagnostic markers in six public whole-genome nanopore runs. The archived species and run accession are shown for each row. Filled circles indicate the species, with numbers showing the read count; open circles indicate marker-spanning molecules that could not be resolved between species, and gray dots indicate no marker-spanning evidence. (B) Marker-spanning reads identified in the biopsy cohort. Specimens A–O are PCR-positive, and the PCR-negative specimen is shown separately. Circle color indicates the best-matching species, numbers show the independent-read count, and empty cells indicate no qualifying reads.

Application of the marker workflow to the clinical dataset recovered 22 qualifying read–locus observations from seven of the 15 PCR-positive biopsies and none from the PCR-negative biopsy (Figure 5B). Biopsy K contained eight concordant *L. tropica* observations distributed across three loci: one *ITS1* read, five miniexon reads, and two cytochrome-*b* reads. This multi-locus marker evidence contrasted with the predominantly *L. aethiopica* genome-wide assignment. Importantly, its direction was consistent with the held-out benchmark, in which authentic *L. tropica* sequence was systematically misassigned to *L. aethiopica* by both genome-wide classifiers. Biopsy K therefore had multiple lines of *L. tropica* diagnostic marker evidence, with no evidence to support the *L. aethiopica* classification.

Biopsy L remained unresolved. It contained only two marker observations: one *L. tropica*-like ITS2 read and one *L. major*-like *HSP70*-coding read. Given the small number of molecules and the limited discriminatory power of *HSP70* coding sequence, this discordance is likely technical, due to low coverage, rather than proof of coinfection or hybrid ancestry.

Overall, marker-spanning reads provided a useful reference-aware check on the unexpected genome-wide classifications. However, their sparse recovery across the cohort and variable discriminatory power prevented marker analysis from supplying definitive species assignments for every specimen.

### 2.6 Multiplexing reduces sequencing costs, but PCR remains cheaper

Because endpoint PCR remains one of the most widely used molecular approaches for detecting *Leishmania*, analytical performance alone is insufficient to determine whether direct sequencing could be adopted by clinical or public health laboratories [3]. Nanopore sequencing is inherently more expensive than a targeted endpoint PCR assay, but the practically relevant questions are how large this cost premium is and how substantially it can be reduced by multiplexing and platform scale. We therefore modeled the per-specimen consumable cost of nanopore sequencing across different batch sizes and compared it with a fully loaded endpoint ITS-PCR batch (Supplementary Figure S2). The purpose was not to perform a formal cost-effectiveness analysis, but to place the additional information generated by sequencing within a realistic economic context.

The modeled sequencing cost declined substantially as fixed library and flow-cell costs were distributed across more specimens. Under an optimized full-use configuration, a 16-specimen MinION run cost approximately US$72.35 per specimen. Scaling the same design to the nominal channel capacity of a PromethION flow cell produced an illustrative batch size of 84 specimens and reduced the modeled cost to approximately US$30 per specimen. By comparison, a fully loaded 24-specimen endpoint-PCR batch cost approximately US$5 per specimen. At these representative batch sizes, the MinION and PromethION scenarios therefore remained approximately 14-fold and 6-fold more expensive than endpoint PCR, respectively.

This comparison is not entirely like-for-like. Endpoint PCR provides an inexpensive answer about the presence or absence of a predefined target, whereas direct sequencing can recover multi-locus parasite evidence and potentially provide species-informative and genome-wide data. Such information could support molecular surveillance, reveal unexpected parasite diversity, and permit retrospective reanalysis as reference databases improve. The present results also demonstrate, however, that these additional benefits are not automatic: species-level interpretation remains dependent on the completeness and representativeness of available reference sequences.

Thus, greater multiplexing and larger flow cells substantially reduced the per-specimen sequencing cost, but nanopore sequencing did not become cost-competitive with endpoint PCR when used solely for routine detection. Its economic justification is more likely to arise in settings where additional genomic information has clinical, epidemiological, or surveillance value beyond a binary PCR result. The estimates should nevertheless be interpreted as consumable-cost scenarios rather than total clinical costs. They excluded DNA extraction, labor, instrument acquisition and maintenance, storage, additional computing, tax, shipping, failed runs, and unused kit capacity. A formal cost-effectiveness evaluation would require these end-to-end costs together with evidence that the additional sequence information improves clinical or public-health decisions.

## 3 Discussion

This study establishes analytical feasibility for recovering genome-wide *Leishmania* evidence directly from biopsy-DNA extracts by PCR-free nanopore sequencing. A deliberately conservative rule requiring three nuclear reads from distinct loci and chromosomes agreed with the PCR comparator in 14 of 15 positive extracts, and qualifying evidence accumulated within minutes to hours of sequence acquisition. Equally important, the experiments define two limits that would be obscured by reporting relative enrichment or classifier agreement alone: adaptive enrichment increased target fraction without increasing absolute target recovery, and two genome-wide classifiers converged on species labels that the held-out benchmark showed to be reference-dependent.

To our knowledge, this is the first evaluation of PCR-free nanopore sequencing for direct detection of *Leishmania* DNA from cutaneous biopsy extracts. Previous nanopore approaches have provided useful detection or typing from amplified products [17–20], while hybrid capture and selective whole-genome amplification have enabled much deeper parasite genomic analysis directly from clinical material [21–23]. The present approach instead asks what can be learned without a parasite-specific wet-laboratory amplification or capture step. That design eliminates primer bias and leaves the complete read available for competitive taxonomic analysis, but it also exposes the workflow to the full host-background and DNA-input constraints of the original extract.

The 93.3% positive percent agreement is encouraging but is not a clinical sensitivity estimate. PCR status was the comparator rather than an independent case definition, the cohort was small, and the single comparator-negative specimen cannot characterize specificity or false-positive behavior. The missed PCR-positive specimen contained one qualifying read, suggesting that it lay below the conservative multi-locus threshold in this run, but no inference about parasite burden can be made without quantitative PCR values, extraction controls, and input-quality data. Prospective validation should include consecutive suspected cases, adequate PCR-negative and alternative-diagnosis controls, blinded analysis, prespecified thresholds, extraction and library blanks, replicate runs, and adjudication against a composite clinical reference standard.

The adaptive-sampling results illustrate why enrichment should be reported in both relative and absolute terms. Adaptive target selection increased the proportion of *Leishmania* bases by as much as 5.8-fold, and the barcode-independent decision audit confirmed strong, although imperfect, rejection of human-classified molecules and retention of qualifying target reads. Nevertheless, the enrichment run produced less total sequence, recovered fewer target bases, and did not consistently shorten time to threshold crossing. Similar dependencies on fragment length, pore utilization, and total yield have been described in metagenomic adaptive-sampling evaluations [26, 27]. Adaptive sampling has successfully supported direct parasite sequencing in other settings [32], but recent clinical-tissue and sputum studies likewise found that compositional gains could be modest or fail to change diagnostic output [28, 29].

The held-out experiment provides a practical warning for metagenomic species reporting. Both alignment and *k*-mer classification assigned most fragments from two *L. tropica* assemblies to *L. aethiopica*, and most *L. donovani* fragments to *L. infantum*. These results do not show that the species are inherently inseparable; they show that the reference panels were non-identifying for these genomes. *Leishmania* genomes are highly syntenic, while species complexes contain substantial population structure, copy-number variation, and genetic exchange [12, 13, 15, 16, 31, 33]. Classifier concordance is therefore weak evidence when the methods interrogate references with the same taxonomic gaps. Future panels should include multiple geographically representative, complete assemblies per species, and benchmark both call accuracy and abstention on held-out populations.

Marker-spanning reads added interpretable but sparse evidence. Established loci differ in copy number, evolutionary rate, and discriminatory range [11]. The concordant *L. tropica*-like ITS1, miniexon, and cytochrome *b* observations in Biopsy K offer a coherent alternative to its genome-wide *L. aethiopica* majority, consistent with the direction of the held-out classifier error. The two discordant observations in Biopsy L are even less conclusive. Cytochrome *b* also represents maxicircle ancestry and should not be treated as an independent nuclear marker in parasites with hybrid ancestry [34]. A reference-aware reporting framework should therefore distinguish genus detection, whole-genome similarity, nuclear marker support, and mitochondrial lineage rather than collapsing them into a single species label.

The apparent speed of sequence acquisition should also be separated from clinical turnaround. The reported minutes-to-hours values exclude DNA extraction, library preparation, computational processing, review, and reporting. The World Health Organization (WHO) target product profile for a point-of-care test for dermal leishmaniases calls for performance and operational characteristics that include high sensitivity and specificity, a result in less than one hour, and low cost [35].

Despite these limitations, the same properties that make direct nanopore sequencing inefficient as a single-target test may make it valuable as a broader diagnostic tool. Unlike a Leishmania-specific PCR, amplification-free long-read sequencing does not pre-specify the organism and could recover evidence from bacterial, fungal, or other parasitic causes of cutaneous lesions with overlapping clinical phenotypes, as well as co-infections. Long molecules provide contiguous taxonomic information, link discriminatory variants within individual reads, and allow the same dataset to be re-analysed as reference databases improve. These benefits were not evaluated directly here and will depend on adequate depth, contamination controls, and representative databases, but they support a potential role in unresolved cases and reference-laboratory investigations where etiological breadth and genomic context may justify the additional cost. That economic balance is also likely to improve as flow-cell yield advance, reducing the cost per informative read and per specimen even if targeted PCR remains cheaper for routine detection alone.

In conclusion, PCR-free nanopore sequencing recovered conservative, multi-locus *Leishmania* evidence directly from most PCR-positive biopsy extracts in this feasibility series. The same dataset shows why target fraction should not substitute for absolute recovery and why agreement between classifiers should not substitute for representative references. Prospective diagnostic validation and population-aware databases are the next requirements for translating direct long-read sequencing into clinically reliable detection and species reporting.

## 4 Materials and methods

### 4.1 Study design, biopsy specimens, and PCR comparator

This analytical feasibility study utilized 16 archived DNA extracts obtained from cutaneous biopsy specimens collected at the referral leishmaniasis clinic of Al-Yahya Primary Health Care Center, Al-Ahsa, Eastern Saudi Arabia. Of these, 15 specimens were positive and one was negative by a PCR assay targeting the ribosomal internal transcribed spacer (ITS) region. Specimens are denoted A–O for the PCR-positive extracts and “PCR-negative” for the comparator negative extract.

Skin samples were obtained by punch biopsy (Kai Industries Co., Ltd., Japan) from patients with a clinical diagnosis of cutaneous leishmaniasis (CL). Clinical suspicion of CL was based on the presence of ulcerated lesions or infiltrative erythematous nodules on exposed body areas in patients residing in Al-Ahsa with a documented history of sandfly bite exposure. The detailed methodology of the primary study has been described previously [30].

Genomic DNA was extracted from skin specimens using a commercial extraction kit (QIAamp Fast DNA Tissue Kit, Qiagen, Germany), according to the manufacturer’s instructions. All samples were processed using a modified protocol targeting kinetoplast DNA (kDNA) as an initial screening step, followed by the protocol described by Schönian et al., which employs two genus- and species-specific PCR assays. Each PCR reaction was performed in a total volume of 25 uL, comprising 12.5 uL of master mix, 1 uL of each primer, 7.5 uL of RNase-free water, and 3 uL of sample DNA (approximately 25 ng). Leishmania major (MHOM/TM/82/Lev) and Leishmania tropica (MHOM/SU/80/K28) reference strains were used as positive controls, and RNase-free water served as the negative control. All PCR reagents were obtained from Promega (USA), and primers were synthesized by Macrogen (Seoul, South Korea). Amplified products were analyzed by electrophoresis on a 1% agarose gel at 120 V in 1× Tris-acetate-EDTA (TAE) buffer and visualized under ultraviolet light.

Species identification was performed by ITS1 PCR/nested PCR-restriction fragment length polymorphism (RFLP) analysis using the HaeIII restriction enzyme (Molequle-On, New Zealand). ITS1 PCR/nested PCR products were bidirectionally sequenced by Macrogen (South Korea). The resulting sequences were deposited in the GenBank database under accession numbers OK560721–OK560817.

### 4.2 Ethical approval

The study protocol and collection of clinical specimens were reviewed and approved by the Institutional Review Board (IRB) of Imam Abdulrahman Bin Faisal University (IRB-PGS-2020-01-427/IRB-PGS-2020-01-186) and the Ministry of Health (MoH) IRB Committee (KFHH RCA:08-25-2020). Written informed consent was obtained from all participants prior to specimen collection.

Clinical procedures and specimen handling were conducted at Al-Yahya Primary Health Care Center, whereas DNA extraction, PCR, post-PCR processing, and nanopore sequencing runs were performed exclusively at Imam Abdulrahman Bin Faisal University. Computational and bioinformatic analyses performed at King Abdullah University of Science and Technology (KAUST) utilized exclusively de-identified sequence datasets containing no personal health identifiers. No human biological specimens, clinical material, or DNA samples were transferred to, stored at, or processed within KAUST laboratories.

### 4.3 Nanopore library preparation and run design

Libraries were prepared without DNA fragmentation or target amplification, following Oxford Nanopore ligation-sequencing protocol for the Native Barcoding. For the 16-plex library, 400 ng of genomic DNA was used per extract; for each three-extract library, 1 *µ*g was used per extract, following the protocol recommendations. DNA underwent combined repair and end repair/dA-tailing with the NEBNext FFPE DNA Repair Mix (M6630) and NEBNext Ultra II End Repair/dA-Tailing Module (E7546; New England Biolabs), followed by AMPure XP bead purification. Each sample was ligated to a unique native barcode using the NEB Blunt/TA Ligase Master Mix (M0367) and ligated to the Oxford Nanopore Native Adapter using the NEBNext Quick Ligation Module (E6056). For each run, 35–50 fmol of the final pooled library in 12 *µ*l was combined with 37.5 *µ*l Sequencing Buffer and 25.5 *µ*l Library Beads, and the complete 75 *µ*l mixture was loaded onto an R10.4.1 MinION flow cell (FLO-MIN114). Sequencing during the 16-plex sequencing run used adaptive human-depletion mode in the minKNOW app interface using the human genome (GRCh38) reference. Subsets of three extracts were additionally sequenced on separate flow cells under a standard mode, in which all molecules were accepted, and an adaptive target-enrichment mode, in which molecules matching a combined *L. major* /*L. tropica* target reference panel were retained and nonmatching molecules were ejected.

### 4.4 Basecalling, demultiplexing, and primary read filters

Raw signal data were rebasecalled with Dorado version 1.3.2 using the super-accuracy model dna_r10.4.1_e8.2_400bps_sup@v5.2.0. Reads were demultiplexed with Dorado using the run-specific SQK-NBD114-24 configuration without barcode trimming. Reads were assigned to specimens only when the barcode was detected at both ends, then retained if their mean Phred quality was at least 10 and their length was at least 500 bp. Reads surviving these steps are termed primary-filter reads. Summary yield was accumulated from primary-filter bases acquired during the first 24 h of each run.

### 4.5 Competitive alignment and the specimen-positive rule

Complete reads were competitively aligned with minimap2 version 2.24 [36], using the map-ont preset, against a composite database containing nuclear *Leishmania* assemblies, two human reference assemblies—GRCh38.p14 (GCF_000001405.40) and T2T-CHM13v2.0 (GCF_009914755.1)—related trypanosomatids, and likely laboratory and environmental contaminants.

A qualifying nuclear *Leishmania* read was required to have at least 80% query coverage, mapping quality (MAPQ) at least 20, aligned span at least 500 bp, nucleotide identity at least 85%, and a positive alignment-score margin between the best *Leishmania*-genus placement and the next-best genus. Nuclear reference coordinates were divided into 10-kb loci. A sequencing-positive specimen required at least three qualifying reads assigned to three distinct, non-overlapping 10-kb loci on at least two chromosomes.

### 4.6 Sequencing-only time to detection

Primary-filter reads were replayed in acquisition order. The first time at which a specimen satisfied all read, locus, and chromosome components of the frozen rule was recorded as sequencing-only time to detection. No specimen preparation or computational/reporting latency was added.

### 4.7 Adaptive-sampling analysis

The stringent dual-ended barcode rule was retained for clinical detection and the other specimen-level analyses. For the matched adaptive-performance comparison, the standard and target-enrichment runs were demultiplexed separately using one-end barcode assignment, because adaptive ejection can terminate a read before the distal barcode is sequenced. Reads assigned to the three paired specimens were then subjected to the same mean-Q-score (at least 10), read-length (at least 500 bp), competitive-alignment, and qualifying nuclear-*Leishmania* rules used in the primary analysis.

For each paired specimen, qualifying nuclear *Leishmania* bases accumulated during the first 24 h were divided by all one-end-assigned primary-filter bases for that specimen to obtain the target fraction. Absolute target recovery was the total number of qualifying nuclear bases in the same interval. Fold enrichment was calculated as the enrichment-run target fraction divided by its standard-run counterpart; absolute recovery was calculated as target bases under enrichment divided by those under standard sequencing.

Terminal adaptive-sampling performance was evaluated without barcode selection. Every read in the deduplicated basecall BAM for the target-enrichment and human-depletion runs was competitively aligned to the composite reference, and the resulting offline origin was linked to the terminal adaptive decision by read ID. Human origin was defined by the best competitive alignment; nuclear target reads were required to pass the complete qualifying-read rule. unblock was summarized as ejected, whereas stop_receiving and no_decision were combined as continued. Reads without a matching terminal-decision record were labeled no log.

Adaptive sampling was performed in MinKNOW version 5.5.5 using real-time Fast-model basecalling. For target enrichment, the reference FASTA contained 79 nuclear sequences: all 36 chromosomes from *L. major* strain Friedlin (GCA_000002725.2) and 43 chromosome or unplaced sequences from *L. tropica* strain CDC216-162 (GCA_014139745.1). Molecules matching either assembly were retained for continued sequencing, whereas nonmatching molecules were ejected. For human depletion, GRCh38 (GCA_000001405.15) was used as the reference; matching human molecules were ejected and nonmatching molecules were retained. The real-time Fast-model calls were used only to control molecule acceptance or ejection; all reported sequence analyses used the offline Dorado SUP basecalls described above.

### 4.8 Genome-wide *k*-mer classification

Kraken2 version 2.17.1 [37] was applied using exact 35-mer evidence and lowest-common-ancestor assignment against a custom species-balanced genomic database containing one reference per included species. Clinical species compositions were released only for specimens meeting the genus-level rule and were normalized among the *L. major*, *L. tropica*, and *L. aethiopica* calls displayed.

### 4.9 Held-out-assembly benchmark

Five nuclear assemblies absent from the classifier databases were divided deterministically into 3-kb fragments sampled at 10-kb intervals. The panel comprised *L. major* JRCGR Lm01, *L. tropica* MHOM/LB/2017/IK (BioProject PRJNA438080; assembly QBEU00000000.1), *L. tropica* MHOM/LB/2015/IK (BioProject PRJNA453461; assembly QEHO00000000.1), *L. aethiopica* L147, and *L. donovani* HU3.

Fragments were processed by the same competitive-alignment and Kraken2 species-assignment workflows. Percentages were calculated among fragments receiving a species-level call. Because the fragments preserved assembly sequence without simulated nanopore error, the benchmark assessed reference-panel representation and classifier separability rather than sequencing-error tolerance.

### 4.10 Marker-spanning read analysis

Seven loci were evaluated: *ITS1*, *ITS2*, miniexon, *HSP20*, *HSP70* coding sequence, *HSP70* 3*^′^* UTR, and maxicircle cytochrome *b*. Candidate spanning reads were discovered with minimap2, compared by BLASTN [38] against all available same-marker accessions in the reference panel, and resolved only when the best-versus-second-species score margin satisfied the prespecified criterion. Each molecule was counted once per locus.

The workflow was first applied to six independent public whole-genome nanopore runs: *L. mexicana* SRR20123517, *L. infantum* SRR21601459, *L. donovani* SRR22035125, *L. panamensis* SRR30096282, *L. guyanensis* SRR30096286, and *L. braziliensis* SRR30096287. For each run–marker cell, the plurality species and its supporting molecule count were reported; unresolved marker-spanning molecules were retained as abstentions. The *L. mexicana* HSP70 3*^′^*-UTR cell used the only complete reference available from the same strain and was excluded from independent accuracy summaries.

### 4.11 Marker-reference divergence

Marker references were aligned with MAFFT version 7.520 [39]: E-INS-i for ITS1, ITS2, miniexon, and HSP70 3*^′^* UTR, and G-INS-i for HSP20, HSP70 coding sequence, and cytochrome *b*. Pairwise differences were counted as substitutions plus internal gap-aligned bases over the shared interval; one-sided terminal overhangs were excluded. Comparisons were repeated under the alternative alignment strategy, and differences exceeding two positions or one percentage point were flagged as alignment sensitive. Exact duplicate accessions were collapsed before all cross-species pairs were formed for *L. major*, *L. tropica*, and *L. aethiopica*.

### 4.12 Cost model

Sequencing-consumable costs were distributed across modeled batch sizes using public US list prices accessed 6 August 2026 [40]. The optimized MinION scenario assumed full use of the SQK-NBD114.96 kit at 16 specimens. An illustrative PromethION batch size was scaled by the nominal 2,675/512 channel ratio to 84 specimens. The endpoint ITS-PCR comparison assumed a fully loaded 24-specimen batch with positive and no-template controls and gel readout. Shared extraction was excluded from all modalities. Sequencing estimates also excluded labor, instruments, service, storage, additional compute, tax, shipping, failed runs, and pack expiry.

### 4.13 Statistical analysis

Positive percent agreement was defined as the number of PCR-positive specimens meeting the sequencing-positive rule divided by all PCR-positive specimens. Its two-sided 95% confidence interval was calculated by the Clopper–Pearson exact binomial method [41]. No specificity estimate was calculated because only one PCR-negative specimen was available. Adaptive-sampling, species-composition, marker, timing, and cost results were summarized descriptively; no hypothesis testing was used.

## Declarations

### Ethics approval and consent to participate

Ethical approval for clinical sample collection, diagnostic testing, and sequencing was granted by the Institutional Review Board of Imam Abdulrahman Bin Faisal University (IRB-PGS-2020-01-427/IRB-PGS-2020-01-186) and the Ministry of Health (MoH) IRB Committee (KFHH RCA:08-25-2020). All experimental wet-laboratory procedures were conducted at Imam Abdulrahman Bin Faisal University. Bioinformatic analysis at KAUST was conducted exclusively on de-identified sequencing data and did not involve human tissue or biological samples.

### Consent for publication

Not applicable. This manuscript does not contain any individual personal identifiers, clinical photographs, or direct identifiable patient information.

### Availability of data and materials

Human-filtered Oxford Nanopore reads generated in this study are available in the NCBI SRA under BioProject accession PRJNA1520088. Public datasets analyzed are available under SRA accessions SRR20123517, SRR21601459, SRR22035125, SRR30096282, SRR30096286, and SRR30096287. Custom scripts and analysis pipelines are available at https://github.com/fffamk/leishmania-nanopore.

## Authors’ contributions

F.A. and A.A.R. conceived and designed the study. A.A.R. secured institutional ethical approval, provided clinical resources, and oversaw clinical specimen collection, DNA extraction, and diagnostic PCR testing. F.A. and A.A.R. performed nanopore library preparation and sequencing at Imam Abdulrahman Bin Faisal University. F.A. developed the bioinformatic pipelines, performed all computational and statistical analyses, and wrote the complete original draft of the manuscript. C.F.-J. provided guidance, resources, and project supervision. A.A.R. and C.F.-J. contributed to critical revision and editing of the manuscript. All authors reviewed and approved the final manuscript.

## Data Availability

Human-filtered Oxford Nanopore reads generated in this study are available in the NCBI SRA under BioProject accession PRJNA1520088. Custom scripts and analysis pipelines are available at https://github.com/fffamk/leishmania-nanopore.

## Acknowledgements

We gratefully acknowledge the Center of Excellence for Smart Health (KCSH) and the Division of Biomedical Sciences at King Abdullah University of Science and Technology (KAUST) for supporting this research and providing computational infrastructure. We also thank the clinical and laboratory staff at the College of Medicine, Imam Abdulrahman Bin Faisal University, for facilitating specimen access and diagnostic support. Specifically, we thank Ms. Salma Al-Jaroodi for her help during library prep and sequencing.

## Generative-AI disclosure

During the preparation of this work, the authors used Google Gemini (Gemini 3.7 with Extended Thinking), OpenAI ChatGPT 5.6 and Anthropic Claude 5 Sonnet to assist in drafting custom bioinformatic scripting (Python and Bash) and to improve the grammatical clarity, conciseness, and readability of the manuscript text. AI tools were not used to conceive or design the study, generate data, conduct the biological or clinical investigations, or draw scientific conclusions. The authors reviewed and edited all AI-assisted outputs and take full responsibility for the scientific integrity and content of the published article.

---

**Figure S1:**
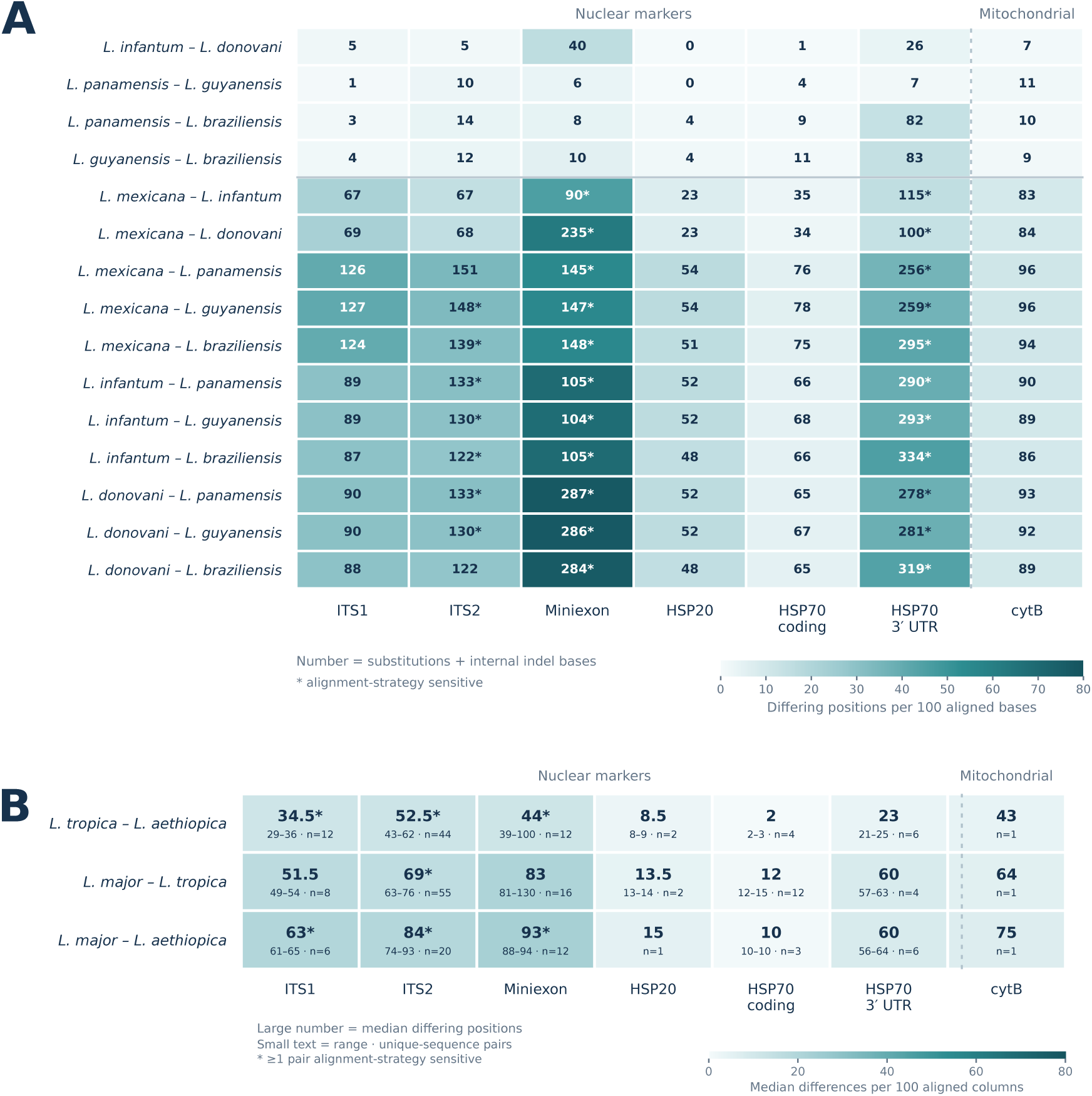
Pairwise divergence among diagnostic-marker reference sequences. (A) Pairwise divergence between the selected marker references for the six species included in the public-read benchmark. Rows represent the 15 possible species pairs. Cell values give the number of substitutions and internal gap-aligned positions within the shared aligned region, while color indicates the corresponding number per 100 comparable positions. Asterisks mark comparisons sensitive to the alignment strategy. Miniexon values should be considered approximate because fragment boundaries and repeat structure affected some alignments. (B) Divergence among all distinct published marker sequences for *L. major*, *L. tropica*, and *L. aethiopica* in the reference panels. Large values show the median number of differing positions; smaller text shows the observed range and number of sequence pairs. Color indicates median length-normalized divergence using the same scale as in A.

**Figure S2:**
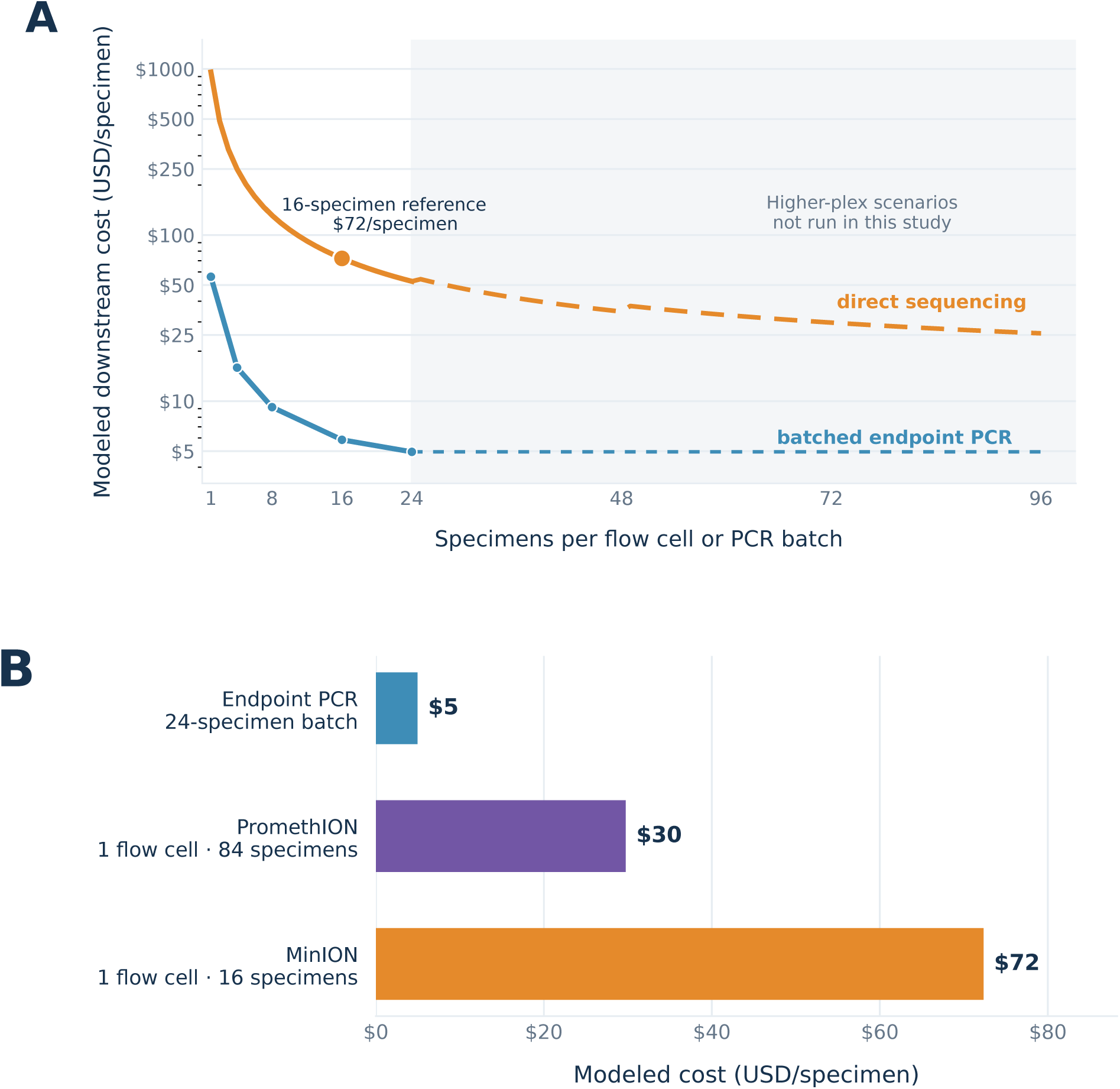
Modeled per-specimen costs for nanopore sequencing and endpoint PCR. (A) Estimated post-extraction consumable cost per specimen across increasing batch sizes. The orange curve represents MinION sequencing with an optimized SQK-NBD114.96 allocation; the filled circle marks the 16-specimen multiplexing level used in this study, and the dashed segment indicates higher multiplexing levels that were not tested experimentally. Blue points represent fully loaded endpoint ITS-PCR batches, including positive and no-template controls and gel-based readout. (B) Comparison of optimized 16-specimen MinION, modeled 84-specimen PromethION, and 24-specimen endpoint-PCR configurations. PromethION capacity was extrapolated from the MinION design by relative channel count and was not evaluated experimentally. Estimates use public US list prices accessed 6 August 2026.

